# Mutations that confer resistance to broadly-neutralizing antibodies define HIV-1 variants of transmitting mothers from that of non-transmitting mothers

**DOI:** 10.1101/2021.01.07.21249396

**Authors:** Amit Kumar, Elena E. Giorgi, Joshua J. Tu, David R. Martinez, Joshua Eudailey, Michael Mengual, Manukumar Honnayakanahalli Marichannegowda, Russell Van Dyke, Feng Gao, Sallie R. Permar

## Abstract

Despite considerable reduction of mother-to-child transmission (MTCT) of HIV through use of maternal and infant antiretroviral therapy (ART), over 150,000 infants continue to become infected with HIV annually, falling far short of the World Health Organization goal of reaching <20,000 annual pediatric HIV cases worldwide by 2020. Prior to the widespread use of ART in the setting of pregnancy, over half of infants born to HIV-infected mothers were protected against HIV acquisition. Yet, the role of maternal immune factors in this protection against vertical transmission is still unclear, hampering the development of synergistic strategies to further reduce MTCT. It has been established that infant transmitted/founder (T/F) viruses are often resistant to maternal plasma, yet it is unknown if the neutralization resistance profile of circulating viruses predicts the maternal risk of transmission to her infant. In this study, we amplified HIV-1 envelope genes (*env*) by single genome amplification and produced representative Env variants from plasma of 19 non-transmitting mothers from the U.S. Women Infant Transmission Study (WITS), enrolled in the pre-ART era. Maternal HIV Env variants from non-transmitting mothers had similar sensitivity to autologous plasma as observed for non-transmitting variants from transmitting mothers. In contrast, infant variants were on average 30% less sensitive to paired plasma neutralization compared to non-transmitted maternal variants from both transmitting and non-transmitting mothers (p=0.015). Importantly, a signature sequence analysis revealed that motifs enriched in *env* sequences from transmitting mothers were associated with broadly neutralizing antibody (bnAb) resistance. Altogether, our findings suggest that circulating maternal virus resistance to bnAb-mediated neutralization, but not autologous plasma neutralization, near the time of delivery, predicts increased MTCT risk. These results caution that enhancement of maternal plasma neutralization through passive or active vaccination during pregnancy could drive the evolution of variants fit for vertical transmission.

**Author Summary:** Despite widespread, effective use of ART among HIV infected pregnant women, new pediatric HIV infections increase by about 150,000 every year. Thus, alternative strategies will be required to reduce MTCT and eliminate pediatric HIV infections. Interestingly, in the absence of ART, less than half of HIV-infected pregnant women will transmit HIV, suggesting natural immune protection of infants from virus acquisition. To understand the impact of maternal plasma autologous virus neutralization responses on MTCT, we compared the plasma and bnAb neutralization sensitivity of the circulating viral population present at the time of delivery in untreated, HIV-infected transmitting and non-transmitting mothers. While there was no significant difference in the ability of transmitting and non-transmitting women to neutralize their own circulating virus strains, specific genetic motifs enriched in variants from transmitting mothers were associated with resistance to bnAbs, suggesting that acquired bnAb resistance is a common feature of vertically-transmitted variants. This work suggests that enhancement of plasma neutralization responses in HIV-infected mothers through passive or active vaccination could further drive selection of variants that couldbe vertically transmitted, and cautions the use of passive bnAbs for HIV-1 prophylaxis or therapy during pregnancy.

## Introduction

Mother-to-child transmission (MTCT) of HIV-1 was responsible for approximately 160,000 new pediatric infections worldwide in 2018 [1], despite wide availability of maternal antiretroviral therapy (ART), which can significantly reduce vertical transmission rates [2]. MTCT occurs via three distinct routes: in utero, peripartum, and postpartum through breastfeeding. Among pregnant women living with HIV-1 not receiving ART, the overall rate of MTCT of HIV-1 is between 30-40% [3]. However, when ART is used during pregnancy, the rate of HIV-1 MTCT can be as low as <2% [4]. Despite the success of ART, factors like limited access and adherence to ART, fetal toxicities [5, 6], development of drug resistant viruses, and acute maternal infection during pregnancy remain to be addressed to further reduce or eliminate pediatric HIV-1 infections [7]. Hence, it is clear that additional strategies will be required to work synergistically with ART to eliminate MTCT of HIV-1.

Interestingly, in the absence of maternal or infant ART, the majority of infants exposed to HIV-1 do not become infected, suggesting a role for natural immunity in protection against MTCT. Yet, the role of maternal immune response in limiting HIV-1 transmission to the infant is still ill-defined. Maternal neutralizing antibodies (nAbs) that are transferred through the placenta to the infant circulation may contribute to this protection. Some studies have suggested that non-transmitting women have higher magnitude of potentially-protective Env-specific IgG responses compared to transmitting mothers [8-10]. Yet, other studies have reported higher levels and breadth of nAbs in women transmitting HIV-1 compared to non-transmitting women [11-14]. These discordant results could be due to small sample sizes, disparate timing of maternal and infant sample collection and route of transmission, and failure to assess the impact of maternal virus variants that have evolved to escape antibody recognition. A better understanding of the role of maternal antibody responses in the context of viral evolution to escape protective antibodies is therefore needed to inform passive or active vaccine strategies that can synergize with ART to eliminate MTCT.

As maternal IgG is transported through the placenta by an active process mediated by placental Fc receptor interactions [15], MTCT constitutes an attractive model for vaccine immunity to understand the role of pre-existing HIV Env-specific IgG in preventing HIV-1 transmission. A previous study from our group demonstrated that infant transmitted/founder (T/F) viruses were significantly more neutralization resistant to paired maternal plasma when compared to non-transmitted maternal plasma viruses [16], confirming the pattern suggested by reports of infant transmitted viruses that are resistant to neutralization by maternal antibodies [17]. To identify correlates of protection in the context of MTCT, several other studies have compared the breadth and levels of nAb in sera of transmitting and non-transmitting mothers using panels of heterologous primary viral isolates of different clades [18-21]. Surprisingly, one recent study identified increased breadth of plasma nAb responses in transmitting versus non-transmitting women, indicating that the breadth of maternal neutralizing responses negatively contributes to MTCT risk [22]. Moreover, we recently described that infant infection can be initiated by an escape variant of broadly neutralizing antibody (bnAb) present in maternal plasma, a finding that raises a potential safety concern for the use of passive bnAbs in pregnancy [23]. However, few studies have been designed to understand the interplay between the maternal antibodies that are transferred to the infant and neutralization sensitivity of the co-circulating viruses on vertical virus transmission, leaving a gap in our understanding of what immune responses should be targeted for maternal immune-based strategies to further reduce MTCT.

To determine whether the profile of autologous plasma and bnAb neutralization sensitivity of circulating viral populations predicts a mother’s risk of vertical virus transmission, we investigated the autologous plasma and bnAb neutralization sensitivity of the circulating viral population from 35 HIV-1 infected women (16 transmitting and 19 non-transmitting) present at or near the time of delivery. Circulating envelope (Env) variants were produced from each mother-infant pair and tested for autologous maternal plasma and bnAb sensitivity. Additionally, we compared the paired maternal plasma sensitivity of non-transmitted HIV Env variants from transmitting and non-transmitting mothers to understand the combined effects of maternal plasma neutralization potency and neutralization sensitivity of circulating viruses on MTCT risk. This detailed understanding of the role of natural antibody responses and potential bnAb therapeutics in pregnant women, and how they could impact viruses transmitted to the infant, is needed to inform the design of immune-based strategies to synergize with ART and to further reduce and eliminate MTCT.

## Results

### HIV-infected transmitting and non-transmitting mother-infant cohort

The U.S.-based Women and Infant Transmission Study (WITS) cohort was utilized in this study to investigate the combined role of maternal antibodies and virus antibody sensitivity on vertical virus transmission. The WITS cohort was enrolled in the early 1990’s, prior to the availability of ART prophylaxis, thereby eliminating the strong impact of ART on MTCT risk and outcome. Two hundred forty-eight study enrollees (83 transmitting and 165 non-transmitting) from the WITS cohort were screened for study inclusion criteria. Selected transmitting mothers met the inclusion criteria of peripartum transmission, defined as follows: infants tested negative for HIV-1 infection at birth by HIV-1 DNA PCR, yet had HIV-1 detectable DNA at one week of age or older [16]. In addition, these HIV-exposed infants were not breastfed [24]. Each peripartum-transmitting mother was then matched with a non-transmitting woman via propensity score matching [25] based on maternal plasma viral load, peripheral CD4+ T cell count at the time point closest to delivery, mode of delivery, and infant gestational age. We selected a total of 35 HIV-1 infected mothers (19 non-transmitting mothers and 16 mother-infant transmitting pairs) with adequate plasma volume available for this study. Ranges for non-transmitting women were 16,360 to 134,325 copies/ml, and 50 to 1045 cell/mm^3^ for viral load and CD4^+^ T cell counts respectively (Table S1). Maternal plasma viral load of the selected transmitting mothers ranged from 4,104 to 368,471 copies/ml, and peripheral blood CD4^+^ T cell counts ranged from 107 to 760 cells/mm^3^ (Table S2).

### Characterization of complete envelope (*env)* gene sequences from transmitting mother-infant pairs and non-transmitting mothers

Single genome amplicons (SGA) for the HIV-1 *env* gene were obtained from the plasma of transmitting mother-infant pairs as described previously [16]. A total of 463 and 465 *env* SGAs were obtained from the mother and infant transmitting pairs, respectively. Additionally, plasma from 19 non-transmitting mothers was used to obtain 645 *env* sequences (Table 1).

**Table 1:** Comparison of samples from Non-transmitting and transmitting mothers.

|  | **Transmitting Mothers** | **Non-transmitting mothers** |
| --- | --- | --- |
| # of Samples | 16 | 19 |
| # of SGAs | 463 | 645 |
| Viral Load (copies/ml) | 87,193 (4104-368471) | 35,784 (16360-134725) |
| CD4+ T cell count (cells/mm<sup>3</sup>) | 413 (107-1049) | 506 (50-1045) |

Neighbor-joining phylogenetic trees were created for the *env* genes to understand the diversity of the viral population present in transmitting mother-infant pairs and non-transmitting mothers at/near time of delivery. A typical highly diverse chronic HIV-1 population was observed in each maternal sample (Fig 1&2), while paired/corresponding infant viral populations were on average less diverse due to recent infection, allowing us to identify 1 or 2 transmitted/founder viruses (T/Fs) at most in each infant (Fig 2). To assess the autologous plasma neutralization sensitivity of the representative *env* variant population circulating in each transmitting and non-transmitting mother, we selected a total of 134 and 146 representative *env* variants, respectively, for Env pseudovirus preparation (5-12 per mother) using an algorithm as described previously [16] (Figs. 1 & 2).

**Fig 1.**
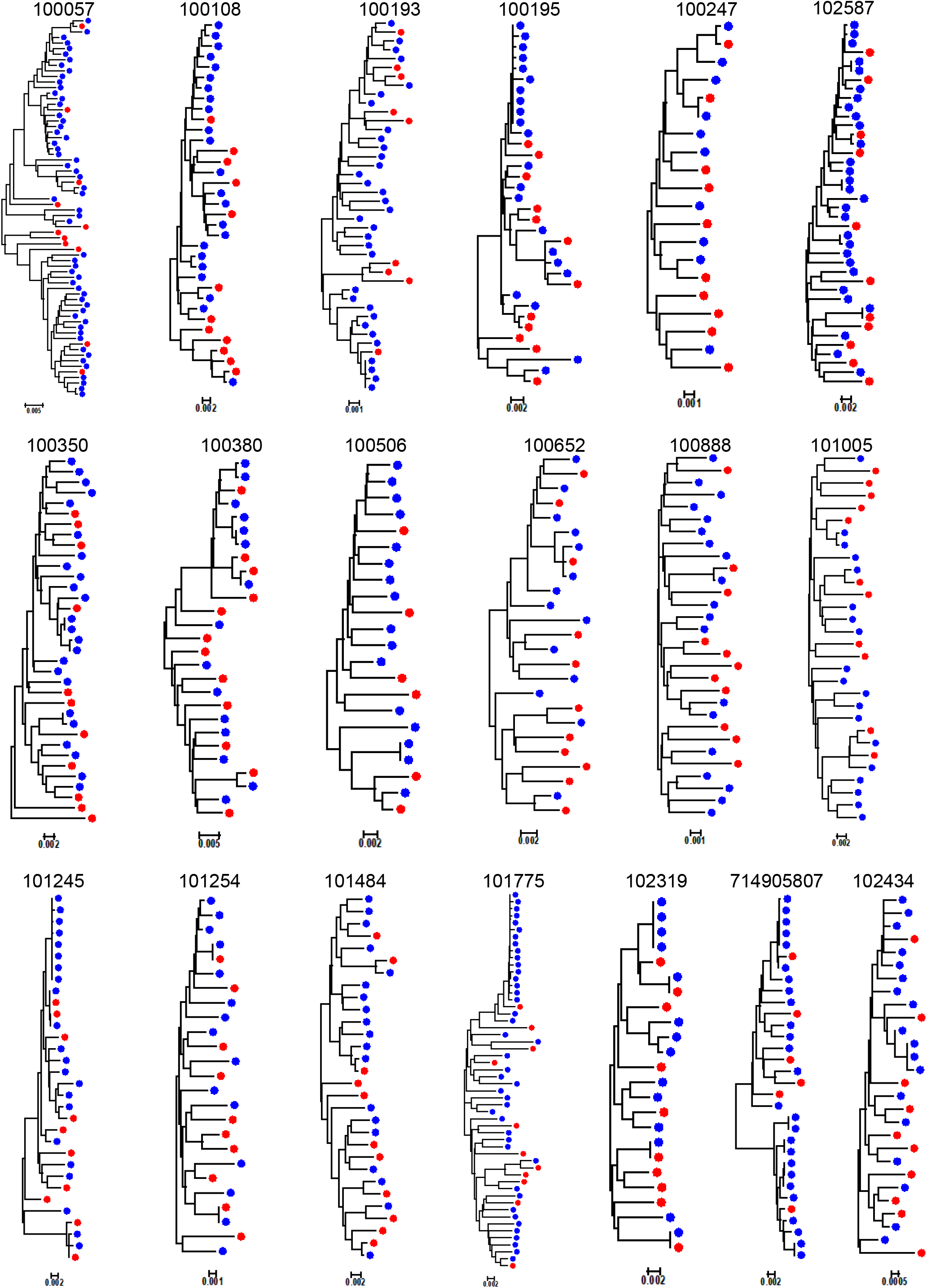
Phylogenetic tree analysis of *env* SGA from non-transmitting mothers. Neighbor joining phylogenetic tree were prepared using the Kimura 2 parameter method. Each colored dot represents an *env* amplicon, while red dots represent the amplicons selected for Env pseudovirus preparation.

**Figure 2:**
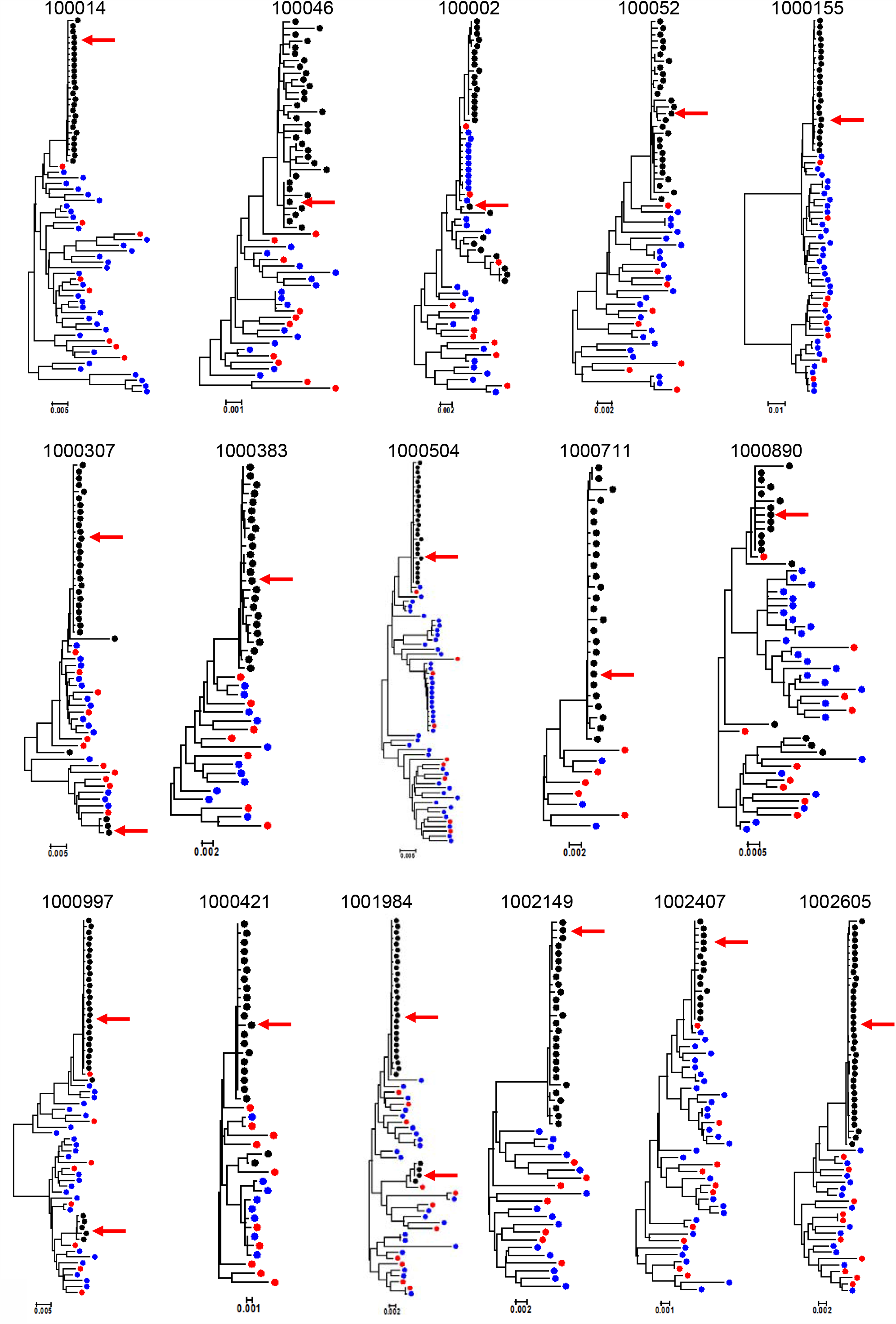
Phylogenetic tree analysis of *env* SGA from transmitting mother-infant pairs. Neighbor joining phylogenetic tree was prepared using the Kimura 2 parameter method. Each infant and mother env amplicons were shown as black and blue dots. Red dots represent the maternal amplicons selected for pseudovirus preparation while infant T/F viruses are shown by red arrow.

### Neutralization sensitivity of circulating HIV-1 Env variants to autologous maternal plasma from peripartum-transmitting and non-transmitting women

HIV Env pseudoviruses were prepared for 280 non-transmitted maternal variants from transmitting (134 viruses) and non-transmitting mothers (146 viruses), along with 19 infant T/Fs to assess their Env neutralization sensitivity to autologous maternal plasma. Variable levels of autologous plasma neutralization sensitivity were observed among non-transmitted *env* variants. In order to test whether the autologous plasma neutralization sensitivity of circulating viruses was significantly different between transmitting and non-transmitting mothers, we fitted a random effect generalized linear model (GLM) with maternal plasma as dependent variable, transmitting status as fixed effect, and maternal ID as random effect. Using the GLM fit, when a predictor was found to be significant (p < 0.05) via ANOVA test between nested models, we proceeded to test the magnitude of the effect using a χ ^2^ test. In addition, differences in number of neutralized viruses were tested using a 2-sided Wilcoxon test. We found no significant difference between transmitting and non-transmitting mothers in the frequency of autologous plasma neutralization-sensitive viruses (p=0.69 by Wilcoxon test) or the autologous plasma neutralization titers against circulating Env variants (p=0.64 by ANOVA test). However, autologous virus neutralization titers against all non-transmitted variants from both non-transmitting and transmitting mothers were on average 1.5-fold higher than infant T/F viruses from transmitting mothers (p=0.005). Furthermore, non-transmitted maternal pseudoviruses were on average 30% more sensitive to maternal autologous plasma than infant T/Fs and their closest viruses in the transmitting mothers (p=0.015) (Fig.3).

**Figure 3:**
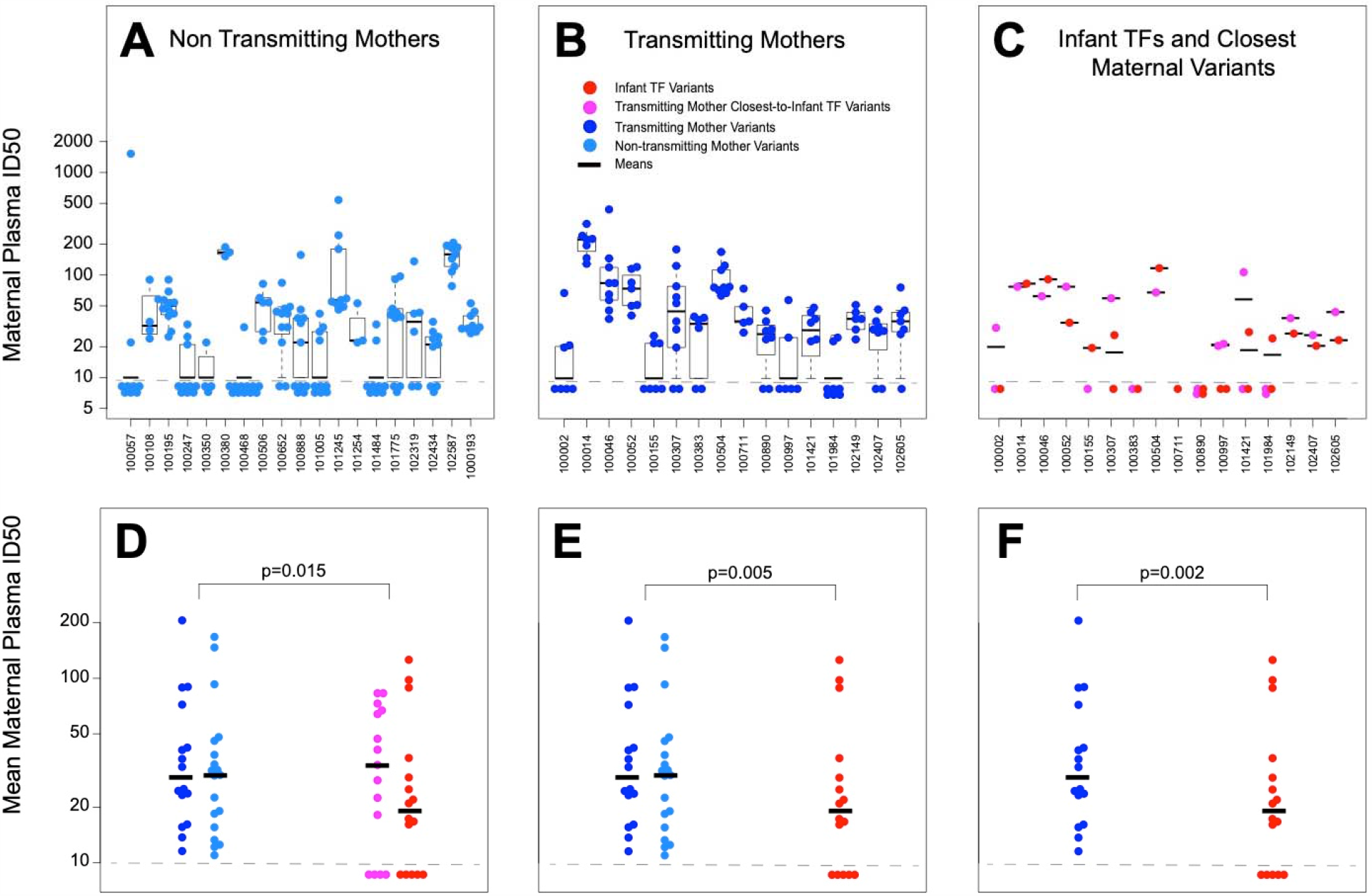
Similar **neutralization sensitivity of non-transmitted maternal variants from transmitting and non-transmitting mothers to paired maternal plasma, while infant T/F variants are more resistant to paired plasma than all maternal variants.** Maternal plasma potency (ID50) against viruses from non-transmitting mothers (A) and transmitting mothers (B) and infant T/Fs (C, red) and their paired closest maternal sequences (C, magenta). Boxes denote interquartile distributions. Bottom panels: geometric mean ID_50_ of non-transmitted variants from each non-transmitting mothers (light blue) and transmitting mothers (dark blue) compared to geometric mean of autologous ID50 of infant T/Fs (red) and closest maternal sequences (magenta). P-values were obtained from fitting a random-effect GLM model. Panel D shows the comparison between all non-transmitted variants and infant T/Fs considered together with the closest maternal sequences. Panel E shows the comparison all non-transmitted variants and the infant T/Fs alone. Panel F shows the comparison between the non-transmitted variants from the transmitting mothers and their paired infant T/Fs.

### Env variant amino acid signatures associated with maternal transmission phenotype and neutralization susceptibility

While we found no differences in magnitude or breadth of plasma autologous virus neutralization responses between transmitting and non-transmitting mothers, we sought to determine if differences in the neutralization sensitivity existed at the viral Env epitope level. Specifically, we looked for particular motifs in Env variants from both transmitting and non-transmitting mothers and infants associated with sensitivity to either autologous plasma, and/or the four second generation HIV-1 bnAbs tested in this study. After excluding infant T/F viruses, we also looked for specific amino acid residues and/or bnAb sensitivity enriched in either transmitting mothers or non-transmitting mothers, and therefore correlated with maternal transmission status. Env pseudoviruses produced from all non-transmitted maternal Env variants (from both transmitting and non-transmitting mothers) and infant T/F were tested against a panel of bnAbs that included PG9 (V2 glycan-specific), VRC01 (CD4bs specific), DH429 (V3 glycan specific) and DH512 (MPER specific) [26-28]. Except for a few exceptions, all tested maternal Env pseudovirus variants were sensitive to neutralization by these four bnAbs (Fig.4).

**Figure 4:**
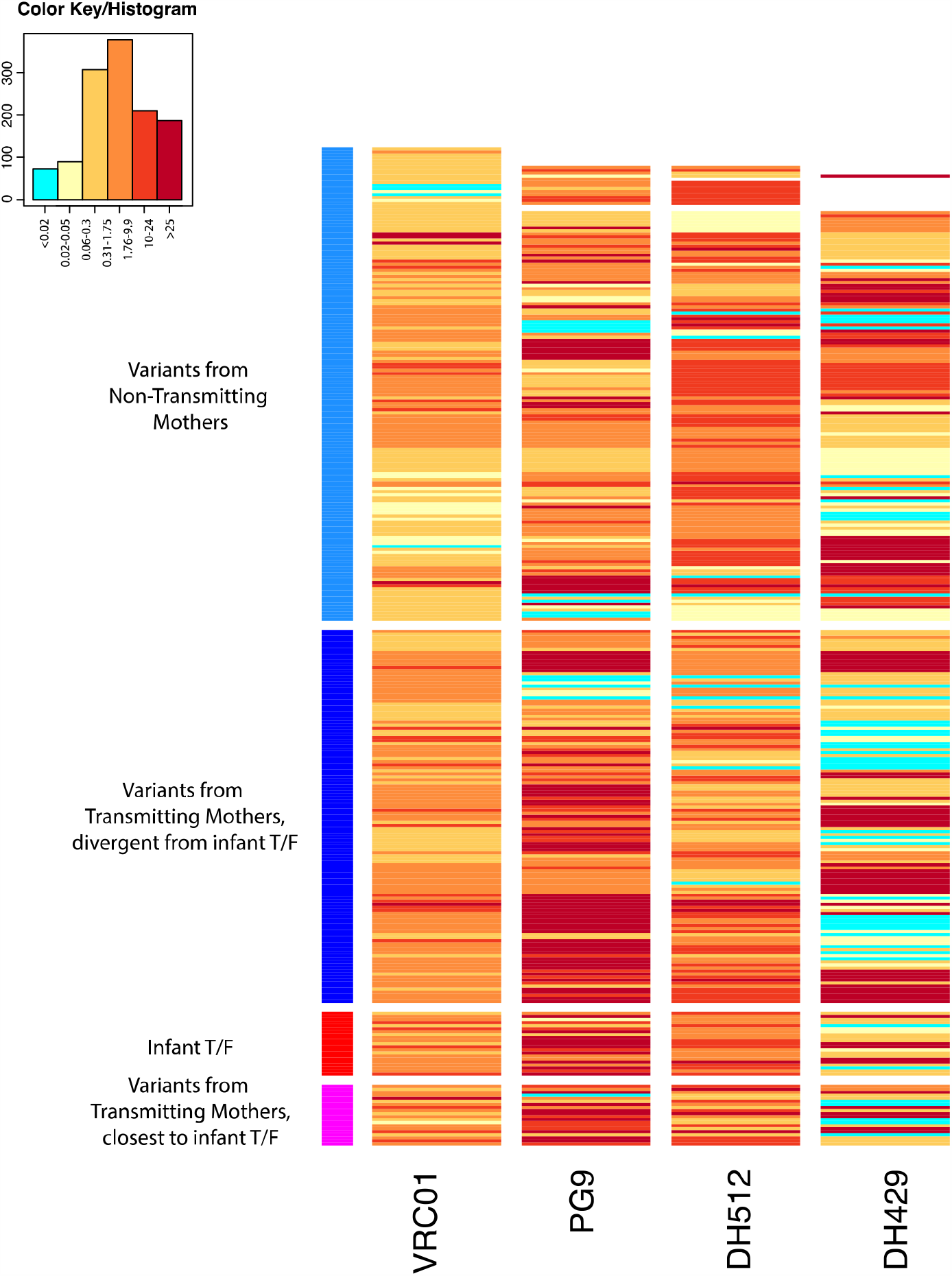
Neutralization sensitivity of infant and maternal Env variants against a panel of bnAbs. Heatmap of bnAbs VRC01, PG9, DH512 and DH429 IC50 against non-transmitting, transmitting, and infant T/F variants, generated using the Heatmap tool on the Los Alamos HIV Database. Rows represent viruses and columns represent bnAbs. The darker hues indicate more potent neutralization, and aquamarine indicates IC50s above threshold, unable to reach this level of neutralization at the highest concentration of bnAb tested. Top rows (indicated by the light blue column to the left) are the variants from the non-transmitting mothers, below (dark blue on the left) are the non-transmitted variants from transmitting mothers, followed by infant T/Fs (red) and transmitting mother variants closest to the infant T/Fs (magenta, bottom).

Using the LANL tool GenSig (https://www.hiv.lanl.gov/content/sequence/GENETICSIGNATURES/gs.html), we performed a phylogenetically corrected signature analysis to identify maternal Env sequences motifs that predicted bnAb sensitivity. The strongest maternal Env amino acid residue associations with bnAb sensitivity were found with sensitivity to PG9 and VRC01. At HXB2 position 234, amino acid N was found to be associated with resistance to VRC01, whereas D was associated with sensitivity (p=1.5×10^−05^ and 1.54×10^−05^ respectively, FDR q=0.00016 and 7.9×10^−05^ respectively; Table 2). Glycosylation site N234 has been implicated as a contact and resistance site in previous studies with CD4bs-specific antibodies [29-32] and with VRC01 in particular, concordant with our findings. In addition, we found site N234 to be strongly associated with maternal transmission status (p=0.0026, FDR =0.02; Table 2), with this PNGS enriched in Env variants from transmitting mothers and the non-PNGS amino acid D enriched in non-transmitting mothers (Fig. 5).

**Table 2:**
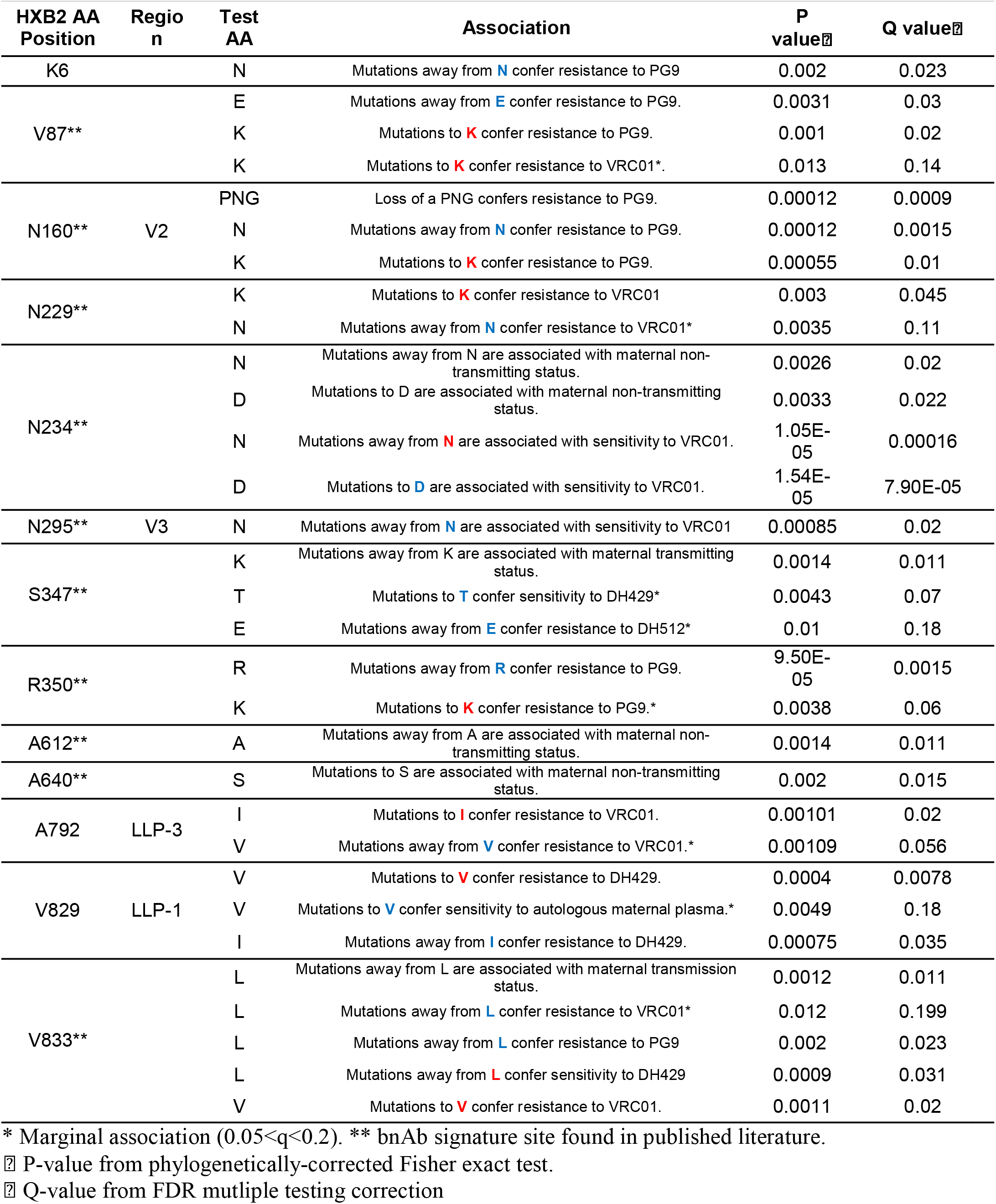
Signature sequence analysis of HIV-1 Env sequence relationship to maternal transmission status, and bnAb sensitivity.

**Figure 5:**
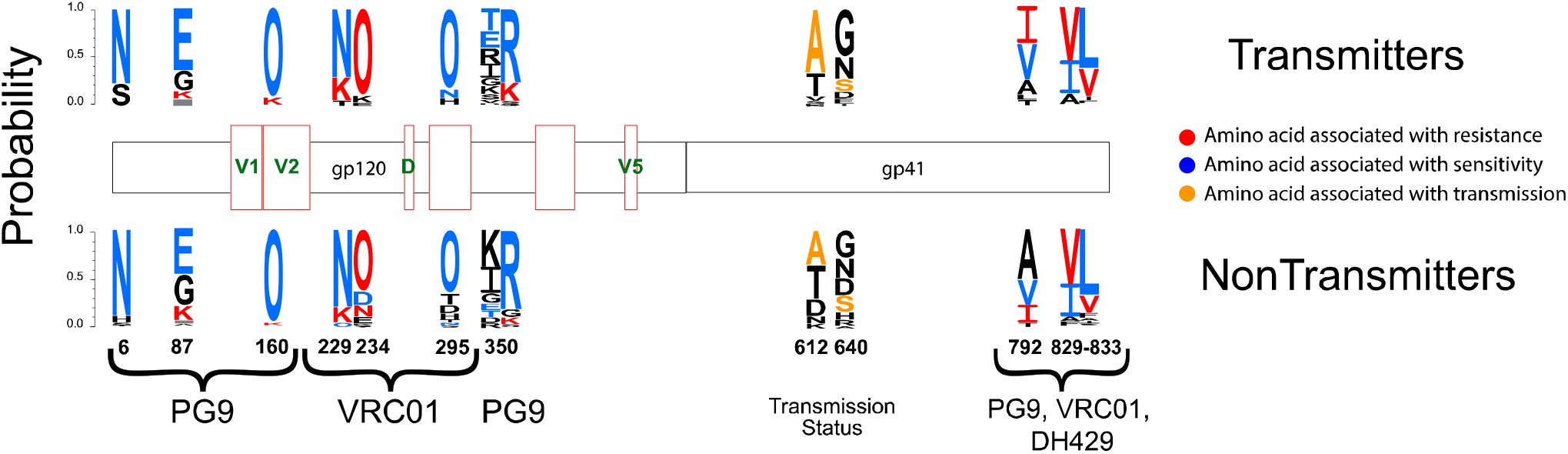
Logo plots of identified signature sequence sites of maternal HIV-1 Env variants that associate with bnAb sensitivity and/or maternal transmission status. Logo plots are shown for each residue that was found to be associated with one of the tested features (see Material and Methods) at the FDR q<0.05 significance level, grouped by transmitting mothers (top) and non-transmitting mothers (bottom). Each amino acid logo is proportional in size to its relative frequency in the alignment. The letter “O” is used to designate N-linked glycosylation sites. Logos colored in red represent mutated amino acids that conferred resistance to one of the bnAbs tested (shown at the bottom), while logos in light blue represent mutations that conferred sensitivity. Orange denotes amino acids associated with transmission status. Notice that while the logo plots represent the frequency of each amino acid, this doesn’t always reflect the counts of ancestral mutations, which is what is tested in the phylogenetically-corrected signature analysis.

Overall, we found 5 Env residues associated with neutralization sensitivity to VRC01, 4 with PG9, and 2 with DH429 at the FDR <0.05 significance level, while no amino acid position was found to be associated with DH429 sensitivity (Table 2, Fig. 5). Eight out of the 11 total sites found in our signature sequence analysis confirmed associations with bnAb sensitivity previously reported by Bricault et al. [31] (Table 2). Unique to our analysis were the findings that amino acids I and V at positions 792 and 829, in the cytoplasmic tail of the Env variants, were associated with resistance to VRC01 and DH429 respectively (Table 2), indicating a conformational change mediated by these mutations that impede CD4bs bnAb recognition. Also, never reported before in the literature was Env site K6, where our analysis showed that mutations away from amino acid N increased resistance to PG9.

All together, our signature sequence analysis found five residues associations with maternal transmission status at FDR <0.05 significance level, of which 3 were also strongly associated with resistance to at least one tested bnAbs (sites N234, S347, and V833, see Table S3) and the remaining two, sites A612 and S640, have been identified by Bricault *et. al* [31) to be associated with increased resistance with V3 glycan-specific bnAbs. Two additional sites, N386 in V4 and S440, both associated with resistance to V3-specific bnAbs [31], were also found to be associated with maternal transmission status, although at a more marginal significance level (FDR =0.08 and 0.07 respectively, Table S3).

As noted above, the amino acid N at site 234 was found to be associated with resistance to VRC01 and, at the same time, a PNG at this site was found to be enriched in Env variants from transmitting mothers. Similarly, at site V833, mutations away from amino acid L were associated with resistance to both PG9 and VRC01 while at the same time being enriched in Env variants from transmitting mothers (Fig 5). Taken together, these findings suggest that acquired resistance to bnAbs may be a factor in selecting for variants that are also fit for vertical transmission, and this bnAb resistance phenotype may be a more important risk factor for transmission than resistance to autologous plasma neutralization.

### Role of HIV-1 Env potential N linked Glycosylation sites (PNGS) in bnAb sensitivity and maternal transmission status

Previous studies have found a higher number of PNGS in V1 to be associated with increased resistance to V3-directed antibodies [31]. While differences in the number of PNGS in the variable regions of Envs from transmitting mothers compared to those from non-transmitting mothers were not large enough to achieve statistical significance in our cohort, a signature sequence analysis identified strong correlations between the loss or acquisition of PNGS in V4 with transmission status (Table 2). Hypervariable regions are extremely hard to align, especially when combining sequences across different donors, which can potentially affect the validity of the results. However, three of the signature sites identified in our analysis as potentially distinct between transmitting and non-transmitting women, N386, N392 and T394, were at the beginning of V4, proximal to the hypervariable portion of the loop (HXB2 positions 396-410), where it is still possible to align sequences across donors (Table 2). The strongest Env PNGs site association with transmission status was at T394, where the acquisition of a PNG was associated with maternal non-transmission status (p=0.0009, FDR =0.0031). At N386, the loss of a PNGS was also associated with maternal non-transmission status (p=0.02, FDR =0.08), whereas the loss of a PNGS at N392 was associated with increased resistance to autologous maternal plasma (p=0.015, FDR =0.06) (Table 2 and S3). This site was the strongest association with maternal plasma sensitivity found among Env sequences in our cohort. Numerous studies have tracked changes in the Env glycan shield as a mechanism for the selection of neutralization escape variants [33, 34] and many glycosylation sites in V4 in particular have been implicated in changes in neutralization sensitivity to V3 and CDbs-bnAbs [31, 35]. Additional studies with more maternal *env* sequences would be warranted in order to establish whether variable loop mutations impacting glycan sites render the maternal viruses more fit for vertical virus transmission by escaping recognition by antibodies that mediate neutralization or other non-neutralizing antibody functions previously reported to be associated with MTCT risk [36-39].

### Broadly neutralizing activity of non-transmitting and transmitting maternal plasma

As our signature analysis found several bnAb resistant sites enriched in variants from transmitting mothers compared to variants from non-transmitting mothers, we wondered whether we could detect broad neutralizing activity from transmitting maternal plasma. We obtained ID_50_ from 15 transmitting and 18 non-transmitting mothers (mother 100888 was excluded due to high murine leukemia virus (MLV) background neutralization activity >60) against a global panel of 9 viruses. We found no statistically significant difference in number of neutralized viruses between the transmitting and non-transmitting mothers (p=0.12 by 2-sided Wilcoxon test). While more non-transmitting mothers (8 out of 18) showed evidence of broad neutralization activity (defined as neutralization of at least four viruses after MLV background subtraction) than transmitting mothers (4 out of 15), this difference was not statistically significant p=0.47 by Fisher exact test) (Fig. 6).

**Figure 6:**
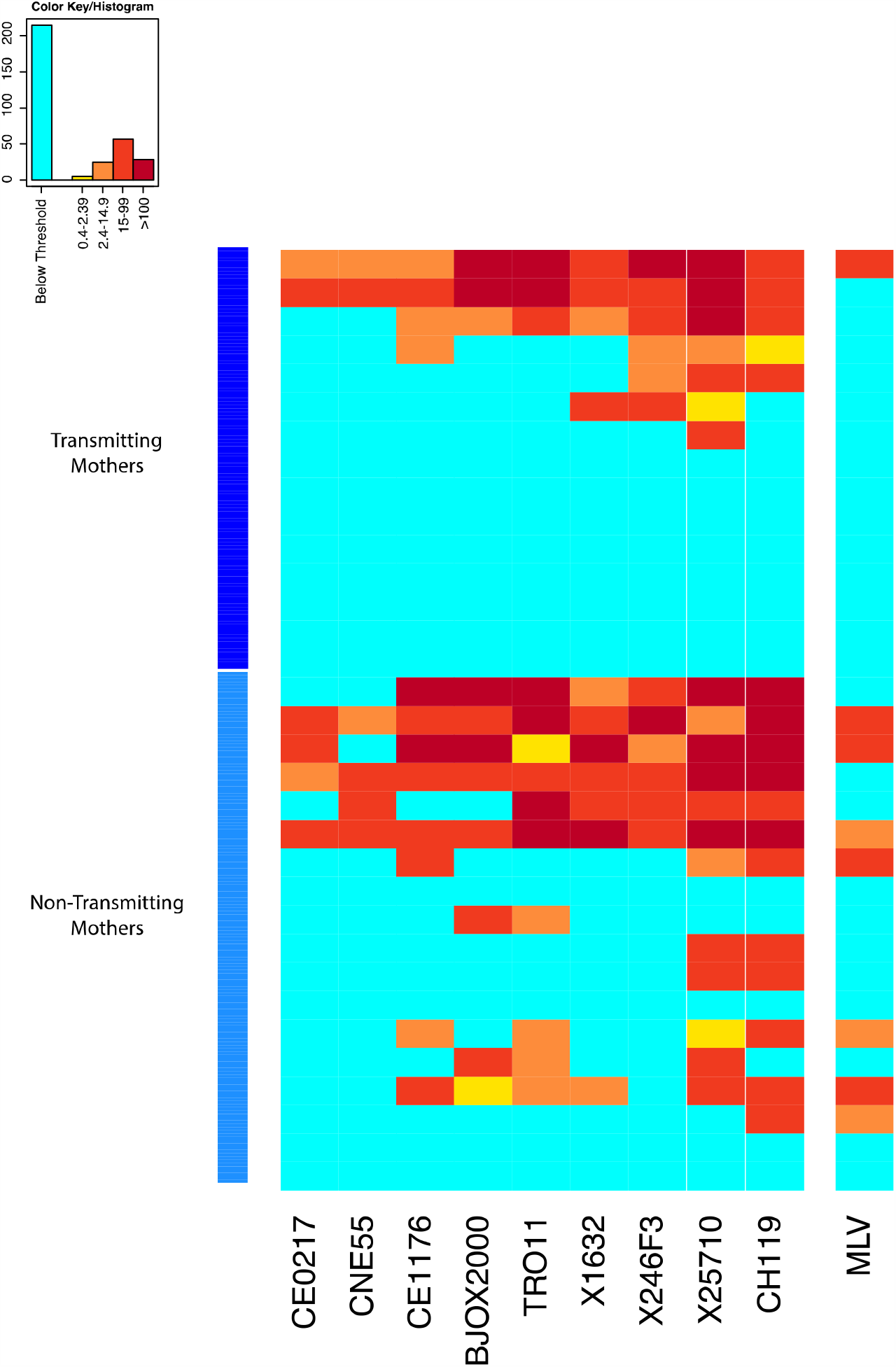
Neutralization sensitivity of global panel of heterologous tier 2 viruses to plasma from transmitting and non-transmitting mothers. Rows represent viruses and columns represent plasma from the transmitting and non-transmitting mothers. The darker hues indicate more potent neutralization, and aquamarine indicates ID_50_s below threshold, unable to reach this level of neutralization at the lowest dilution. Top rows (indicated by the dark blue column to the are the plasma from the transmitting mothers, below (light blue on the left) are plasmas from transmitting mothers.

Since four transmitting mothers (100014, 100504, 102149, and 100307) showed bnAb activity against the panel of global viruses (as defined above), we looked specifically at the infant TFs from those mother-infant pairs and their respective closest maternal sequences to see whether they were enriched for bNAb resistance amino acid at the significant sites found by our signature analysis. These sequences were enriched for bNAb resistant amino acids at 5 out of 9 sites (data not shown). Of note, all sequences had a PNG at site 234, which, as previously noted, is associated with resistance to VRC01. In addition, compared to the closest maternal sequences, the infant TFs were enriched to the resistant-inducing AA V at both positions 829 and 833.

## Discussion

Maternal and infant ARV treatment has significantly reduced the rate of MTCT to low levels, but a maternal or infant vaccine is still needed to eliminate pediatric HIV-1. It has been shown in non-human primate models of sexual transmission that passive immunization of human monoclonal bnAbs that potently neutralize the challenge virus can protect against virus acquisition [40-42]. Considering the established protective role of nAbs and limited success of HIV vaccines that do not elicit bnAbs, it is likely that a vaccine would have to induce bnAbs in order to be highly effective against HIV-1 acquisition [43-46]. MTCT of HIV-1 is a unique setting where the infant receives maternal antibodies generated against autologous viruses to which the infant is exposed from the mother *in utero* [47]. As over half of infants are naturally protected against MTCT, it is believed that protective immune factors, such as maternal antibodies, may prevent the transmission of viruses. Yet, extensive studies of this phenomenon have not firmly established a protective role of maternal antibodies, which may be related to the need to study the interplay between the maternal antibodies and autologous viruses to which the infant is exposed. In this study, we compared the autologous neutralizing antibody responses against the maternal viruses present near the time of delivery in peripartum transmitting and non-transmitting mothers to determine if the ability of the mother to neutralize her own viruses predicts her risk of vertical virus transmission. Despite infant transmitted variants consistently demonstrating autologous plasma neutralization resistance, our analysis revealed no statistically significant differences in the magnitude or frequency of neutralization responses against circulating autologous viruses from transmitting and non-transmitting mothers. Thus, the frequency and potency of maternal neutralization responses against her own circulating viruses do not appear to establish the risk of vertical virus transmission.

We focused on peripartum-transmitting mothers from the WITS cohort who did not receive ART, thereby eliminating the impact of ART on virus selection [24]. Further, to eliminate the clinical factors known to be associated with risk of MTCT, propensity score criteria was used to match non-transmitting mothers with peripartum transmitting mothers which included the CD4+ T cell count, viral load, and mode of delivery. In total, we obtained 1108 Env variants (463 T and 645 NT) from 16 transmitting and 19 non-transmitting mothers. With an average of 30 sequences per mother (range 20-42 sequences), we are 95% confident that these Env variants represented the heterogeneity of each maternal sample present at the time of delivery at a population frequency of 15% or higher. Not many prior studies of the role of maternal antibodies in MTCT have included autologous maternal virus population sequences or functional autologous viruses in their investigations. Other studies have used either partial Env sequences to represent the maternal viral diversity or a small number of transmitting and non-transmitting mother-infant pairs, which limits power to detect sequence diversity among each group [48, 49]. Yet, this study is one of the first to generate autologous single genome Env variants from transmitting and non-transmitting mothers to accurately assess the function of maternal antibodies against co-circulating vertically transmitted and non-transmitted maternal variants.

To investigate our hypothesis that variants from non-transmitting mothers are more sensitive to autologous plasma than in transmitting mothers, Env pseudoviruses were prepared using the SGA *env* sequences from the maternal plasma systematically selected as representative of maternal viral population in order to cover the diversity of the HIV-1 population in each mother. Interestingly, we did not observe statistically significant differences in neutralization sensitivity among viruses between transmitting and non-transmitting mothers. There are only a couple of comparable studies where conflicting results have been observed. Baan et.al [50] compared autologous plasma neutralization sensitivity of viral variants from 7 transmitting and 4 non-transmitting mothers and found that viruses from transmitting mothers were more sensitive to maternal plasma neutralization than the variants from non-transmitting mothers. However, plasma samples used in this study were not contemporaneous, further complicating the analysis. On the other hand, a study by Milligan *et. al* [51] showed that there is no difference in neutralization sensitivity of viral variants from transmitting and non-transmitting mothers as observed in this study. However, the viral variants in Milligan *et*.*al* study were obtained using PBMC DNA which may not represent the circulating variants at the time of transmission. Notably, in the present study, peripartum transmitting and non-transmitting mothers were carefully clinically matched by a propensity score and the plasma employed to isolate viruses was from the delivery time point. Additionally, utilizing a pre-ART era cohort, eliminated viral selection pressure due to ART and further strengthening the results.

When we compared infant T/Fs and their phylogenetically closely related maternal variants with non-transmitted variants from both group of mothers, infant T/Fs and their closely related maternal variants were significantly more resistant to paired maternal plasma. This finding suggests that neutralization resistance to paired maternal plasma is a defining feature of infant transmitted variants. While a few studies [17] have indicated that virus(es) transmitted to infants are neutralization escape variants, other studies contradict these findings [48, 52, 53]. These conflicting results could be due to difference in timing of maternal-infant sample collection, small sample sizes, and undefined routes of transmission. Importantly, we used well-defined criteria for peripartum transmission with a large sample size, adding to the robustness of results. Additionally, isolation of 20-30 SGAs per infant and maternal plasma sample and neutralization testing of 5-12 viruses per mother provide considerably more functional viral sequence data than many previous studies.

Glycosylation sites are known to be targets of numerous bnAbs and hence may have a role in driving the selection of neutralization escape variants in infant infection [33]. However, we did not see any significant differences in number of PNGS and variable loop lengths among infant T/F and non-transmitted maternal variants. In contrast, a few studies have investigated the number of PNGS and variable loop length of infant T/Fs and non-transmitted maternal variants from transmitting mother-infant pairs and found that transmitting viruses had shorter variable loop lengths and less PNGS [48, 49]. Several studies involving chronic HIV transmission in adults where escape variants have been reported to have longer variable loop lengths and more PNGS, indicating that virus escape from maternal antibodies in MTCT may be distinct from that of chronic infection [54-56].

Previously, we found that maternal V3-specific IgG levels and weak (tier 1) neutralization response predicted a reduced risk of transmission in this cohort [21]. In a follow-up study, we showed that infant T/Fs are escape variants of V3 region-specific antibodies [23]. Moreover, it has been shown in rhesus macaque studies that passive immunization by polyclonal or weakly-neutralizing nAbs can reduce vertical transmission risk [57, 58]. So far, there have been two studies in humans involving maternal passive immunizations by polyclonal HIVIG to prevent MTCT and both did not show any additional benefit to ART [59, 60]. These results indicate that targeting specific epitopes of autologous viruses may be required for a successful vaccine to prevent MTCT. Several second generation bnAbs have been isolated from chronic HIV-1 infected patients in recent years and are under study as a passive immunization and/or therapy [61-64]. Hence, it is important to investigate the neutralization efficacy of prototype bnAbs i.e. VRC01 (CD4bs specific), PG9 (V1V2 glycan specific), DH512 (MPER specific) and DH429 (V3 glycan specific) against all Env variants from transmitting and non-transmitting mothers and infant T/Fs to determine what the clinical impact of bnAb prophylaxis and/or therapeutics may be in this setting. Almost all the Env variants tested were sensitive to these bnAbs except few maternal variants. This finding supports the potential efficacy of the ongoing clinical assessment of VRC01 bnAb passive immunization of high risk, HIV-exposed infants as a strategy to further reduce infant HIV acquisition [65]. Also, a comprehensive recent study by Bricault et al. [31] showed the advantages of epitope-based vaccine design based on signature sequence analysis of bnAb-resistant and sensitive variants. Using neutralization epitopes for the bnAbs used in this study were previously defined, a signature sequence analysis was performed to identify amino acids associated with transmission and resistance. We found a number of genetic motifs that were significantly enriched in Env variants from transmitting mothers at positions that were also associated with resistance to V2-, CD4bs-, and MPER-specific bnAbs like PG9, VRC01, and DH512. Interestingly, 3 out of 5 motifs identified in our signature analysis to be significantly associated with maternal transmission status at the FDR <0.05 level, were also associated with increased resistance to either VRC01 or PG9 (Table 2). Importantly, a signature sequence analysis of viral variants from transmitting and non-transmitting mothers demonstrated that certain V1 and V4 loop region amino acids associated with maternal transmission potential, but not necessarily neutralizing sensitivity, which may suggest escape from other maternal antibody functions, such as ADCC, can define the transmission potential of HIV Env variants. Many amino acid positions from region C1, V1V2, C4 and cytoplasmic tail were also associated with viral resistance to the autologous plasma, yet not viral transmission potential. Further, we found 2 glycosylation sites in V4 that associated with neutralization sensitivity, one associated with maternal transmission status and one with resistance to autologous maternal plasma. Interestingly, at the FDR <0.1 significance level, we found 7 signature sites associated with maternal transmission status, of which 3 were also associated with neutralizations sensitivity to PG9, VRC01, and/or DH429, and the remaining four were all sites previously found to be associated with neutralization sensitivity to V3-specific bnAbs [31], suggesting that bnAb-mediated immune pressure may be a major force driving selection of resistant variants in transmitting mothers that are also fit for MTCT transmission.

Since some studies have shown association of nAbs presence or titers against heterologous HIV-1 strains as risk factor for MTCT [11, 12], we also investigated the bnAb activity of the plasma from transmitting and non-transmitting mothers against a reference panel of 9 diverse global HIV-1 viruses. While we did see a trend towards more transmitting than non-transmitting women with pre-defined plasma bnAb activity, defined as neutralization of at least 4 of 9 tier 2 heterologous viruses on the global panel, we did not observe statistically significant difference in the breadth of neutralization activity in the plasma of 16 transmitting and 18 non-transmitting maternal plasma. Our results are in contrast to a study of breast-feeding HIV-1 transmission pairs [22] where transmitting mother showed higher breadth of heterologous virus neutralizing activity compared to non-transmitting mothers. However, different mode of transmission could have led to this difference in results. Also, we did observe higher MLV neutralizing activity among the non-transmitting mothers in this study, potentially indicating distinct sample handling or exposure to ART. Previously, we had shown that infant T/Fs are the escape variants of maternal plasma [16]. Our results along with previous studies indicate that passive or active vaccines inducing bnAb response in pregnant women will need to be used with caution to prevent MTCT.

Overall, this is the largest study comparing plasma neutralization responses against autologous viruses in HIV-infected transmitting and non-transmitting mothers. This unique study design revealed that while infant T/Fs are more likely to be escape variants from maternal neutralization responses when compared with the non-transmitted maternal variants, the mother’s natural ability to neutralize her own circulating viruses does not define a mother’s MTCT risk. Concerningly, we identified that maternal transmission status was tied to Env amino acid signatures that confer resistance to bnAbs, including those being used in clinical trials for HIV-1 prevention and therapy [66, 67] (https://clinicaltrials.gov/ct2/show/NCT03571204). These results caution the use of passive or active vaccine strategies targeting plasma bnAb activity in pregnant women and indicate that more work will need to address the risks of bnAb escape variants that are fit for virus transmission. Moreover, they suggest that there are functional antibody responses other than nAbs, such as ADCC, that may be more important protective factors that define transmitting mothers from non-transmitting mothers. Thus, maternal vaccine strategies to further reduce pediatric HIV infections should be designed to induce multispecific neutralization responses and potentially other non-neutralizing antibody functions that provide potent protection when transferred to the infant, but also eliminate the risk of selecting viruses that can escape maternal antibody functions and become vertically-transmitted variants.

## Data Availability

All data are fully available without restriction. All relevant data are within the manuscript and its Supporting Information files.

## Acknowledgements

We acknowledge Youyi Fong in selecting samples from the WITS cohort, Bette Korber and Kshitij Wagh for useful discussions on the signature analysis. We would also like to acknowledge the support of Pediatric HIV/AIDS Cohort Study (PHACS) team for their management of the Women and Infant Transmission Study cohort repository samples, supported by the *Eunice Kennedy Shriver* National Institute of Child Health and Human Development (HD052102 and HD052104). This work was financially supported by National Institute of Health (NIH) with the RO1 grant number AI122909.

## Materials and Methods

### Study subjects and sample collection

A total of 35 HIV-1 infected women living with HIV-1 (16 transmitting and 19 non-transmitting mothers) were selected from the WITS (Women Infant Transmission Study) cohort based on propensity score and adequate plasma volume (2.0 ml). The WITS cohort was enrolled in the pre-ART era during 1993/1994 in US, consisting of HIV-1 subtype B infections. Propensity score based on established risk factors for MTCT, including maternal CD4^+^ count, plasma viral load, and mode of delivery was used to match non-transmitting women to peripartum-transmitting women [68].

### Ethics statement

Samples used in this study were obtained with informed consent from participants of the Women Infant Transmission Study (WITS) [24]. WITS repository cohort samples were received as de-identified material and were deemed as research not involving human subjects by Duke University Institutional Review Board (IRB). The reference number for that protocol and determination is Pro00016627.

### Viral RNA extraction and SGA analysis

Viral RNA extractions and SGA analyses were done as described previously [16]. Briefly, viral RNA was purified from the plasma sample from each patient by the Qiagen QiaAmp viral RNA mini kit and subjected to cDNA synthesis using 1X reaction buffer, 0.5 mM of each deoxynucleoside triphosphate (dNTP), 5 mM DTT, 2 U/mL RNaseOUT, 10 U/mL of SuperScript III reverse transcription mix (Invitrogen), and 0.25 mM antisense primer 1.R3.B3R (5’-ACTACTTGAAGCACTCAAGGCAAGCT TTATTG-3’). The resulting cDNA was PCR amplified using Platinum Taq DNA polymerase High Fidelity (Invitrogen) so that < 30% of reactions were positive in order to maximize the likelihood of amplification from a single genome. A second round of PCR amplification was conducted using 2μl of the first round products as template. Round 1 amplification conditions were 1 cycle of 94°C for 2 minutes, 35 cycles of 94°C for 15 seconds, 58°C for 30 seconds, and 68°C for 4 minutes, followed by 1 cycle of 68°C for 10 minutes. Round 2 conditions were one cycle of 94°C for 2 minutes, 45 cycles of 94°C for 15 seconds, 58°C for 30 seconds, and 68°C for 4 minutes, followed by 1 cycle of 68°C for 10 minutes. Round 2 PCR amplicons were visualized by agarose gel electrophoresis and sequenced for envelope gene using an ABI3730xl genetic analyzer (Applied Biosystems). Partially overlapping sequences from each amplicon were assembled and edited using Sequencher (Gene Codes, Inc).

### Sequence alignment

All maternal and infant envelope sequences were aligned using the Gene Cutter tool available at the Los Alamos National Laboratory (LANL) website (http://www.hiv.lanl.gov/content/sequence/GENE_CUTTER/cutter.html) and then refined manually. Full-length envelope sequences were manually edited in Seaview [69].

### Pseudovirus preparation

CMV promoter was added to maternal envelope SGAs using the overlapping PCR method and used to prepare pseudoviruses [70]. Briefly, pseudoviruses were prepared by transfection in HEK293T (ATCC, Manassas, VA) cells with 4μg of CMV-env PCR product and 4μg of env-deficient HIV plasmid DNA using the FuGene 6 transfection reagent (Promega) in a T75 flask. Two days after transfection, the culture supernatant containing pseudoviruses was harvested, filtered, aliquoted, and stored at −80°C. An aliquot of frozen pseudovirus was used to measure the infectivity in TZM-bl cells. 20μl of pseudovirus was distributed in duplicate to 96-well flat bottom plates (Co-star). Then, freshly trypsinized TZM-bl cells were added (10,000 cells/well in Dulbecco’s modified Eagle’s medium (DMEM)-10% fetal bovine serum (FBS) containing HEPES and 10μg/ml of DEAE-dextran). After 48 h of incubation at 37°C, 100μl of medium was removed from the wells. 100μl of luciferase reagent was added to each well and incubated at room temperature for 2 min. 100μl of the lysate was transferred to a 96-well black solid plate (Costar), and the luminescence was measured using the Bright-Glo™ luminescence reporter gene assay system (Promega).

### Neutralization assays

Neutralizing antibody activity was measured in 96-well culture plates by using Tat-regulated luciferase (Luc) reporter gene expression to quantify reductions in virus infection in TZM-bl cells. TZM-bl cells were obtained from the NIH AIDS Research and Reference Reagent Program, as contributed by John Kappes and Xiaoyun Wu. Assays were performed with HIV-1 Env-pseudotyped viruses as described previously [71, 72]. Viruses (∼50,000 relative light unit equivalents) were pre-incubated with plasma (starting dilution 1:20) or Mab (starting concentration 25ug/ml) serially diluted 3-fold in a 96 well plate for 1 hr at 37°C before addition of TZM-bl cells. Following a 48-hr incubation, cells were lysed and Luc activity determined using a microtiter plate luminometer and Briteglo (Promega). Neutralization titers are the sample dilution (for serum/plasma) or antibody concentration (monoclonal antibodies) at which relative luminescence units (RLU) were reduced by 50% compared to RLU in virus control wells after subtraction of background RLU in cell control wells. Serum/plasma samples were heat-inactivated at 56°C for 1 hr prior to assay.

### Statistical analyses

Infant T/Fs were identified for each sample as described previously [16, 73]. Briefly, infant envelope sequences were aligned using Seaview and consensus sequence was generated. SGA similar to consensus sequence was used as infant T/F. To select maternal non-transmitted variants and capture the most divergent sequences from the infant T/F, we devised an algorithm as previously described [16]. The algorithm finds the most variable positions in the amino acid alignment and ranks all sequences with respect to the frequencies at these positions. Sequences are then selected starting from the most divergent based on motif coverage as observed in the alignment and in the phylogenetic tree (in other words, if a group of diverging sequences all share the same motif, only one in the group and/or tree node is selected). Differences in number of neutralized viruses were tested using a 2-sided Wilcoxon test. Magnitude of maternal plasma from transmitting and non-transmitting mothers, and between transmitted and non-transmitted variants, were compared using a random-effect generalized linear model (GLM) using maternal plasma as dependent variable, transmitting status as fixed effect, and maternal ID as random effect. Using the GLM fit, when a predictor was found to be significant via ANOVA test between nested models, we proceeded to test the magnitude of the effect using a χ ^2^ test. All tests were conducted on the R platform [74] [http://www.R-project.org]. The GLM was implemented using the lme4 package [75].

Plasma neutralization titers were treated as positive if above the 1:60 dilution threshold. MLV background was subtracted when detected, else a nominal threshold of 60 was subtracted instead. Breadth was measured as number of positive titers after subtracting background as described, and groups were compared using a 2-sided Wilcoxon test (implemented in R).

### Genetic signature analysis

We performed phylogenetically corrected signature analyses to identify amino acid and glycosylation sites associated with transmitting vs. non-transmitting status, maternal plasma, and sensitivity to bNAbs PG9, DH512, DH429 and VRC01. This was done using the LANL tool GenSig [https://www.hiv.lanl.gov/content/sequence/GENETICSIGNATURES/gs.html], which identifies sites of interest using a phylogenetically corrected approach to minimize false positives due to lineage effects [31, 76, 77]. Briefly, at each site, GenSig performs a Fisher exact test of a 2×2 matrix where the rows represent the two states of a feature (i.e. transmitting vs. non-transmitting, or above or below threshold for neutralization sensitivity data), and the columns represent the two possible ancestral states in the phylogenetic tree. For example, if A is the amino acid being tested, then GenSig performs two tests, one where “A” is the ancestral state, and one where “!A” is the ancestral state. For the former, the columns in the Fisher exact matrix will be the counts of how many leaves came from an A->!A transition and how many from A->A respectively, whereas for the latter the counts will be for !A->A and !A->!A respectively. For more details see the GenSig tool explanation (https://www.hiv.lanl.gov/content/sequence/GENETICSIGNATURES/help.html). Because our data consists of multiple sequences from each mother-infant pair, ignoring this phylogenetic correction could potentially yield to spurious associations driven by within-subject correlations. For this analysis, maternal plasma neutralization sensitivity (above or below neutralization threshold of 1:50) and transmission status were treated as dichotomous variables, whereas PG9, DH429, DH512 and VRC01 IC50s were considered multiple ways (upper quartile vs. lower three, lower quartile vs. upper three, and above/below threshold). For robustness, we deemed as viable results only sites that were confirmed with at least two approaches across all considered phenotypes or that had previously been found to be associated with sensitivity in the literature. For multiple testing correction we used a false discovery rate (FDR) of q < 0.2 [78] to screen the results, and then tiered the strength of the associations by q<0.05, q<0.1 and q<0.2 significance levels, the latter being the most marginal findings. Logo plots were created using the LANL tool AnalyzeAlign (https://www.hiv.lanl.gov/content/sequence/ANALYZEALIGN/analyze_align.html).

### bnAb Activity analysis of plasma from transmitting and non-transmitting mothers

Neutralization of 10 viruses incorporating a reference Env from the global panel was tested against a 1:60 plasma dilution from non-transmitting mothers as described previously [79-81]. Pseudovirus prepared with Env glycoprotein from Murine Leukemia Virus (SVA.MLV) was used as a negative control.

## Supporting information

**S1 Table**. Clinical characteristics and number of single genome amplified (SGA) plasma HIV env sequences isolated for non-transmitting mothers.

**S2 Table**. Clinical characteristics for transmitting mothers and their infants.

**S3 Table**. Signature Sequence analysis of the envelope gene and association of amino acid sites with transmission.

